# Prevalence and Burden of Chronic Hepatitis B Virus Infection in Sierra Leone, 1997-2022: Evidence from a 25-Year Systematic Review and Meta-Analysis

**DOI:** 10.1101/2022.11.16.22282393

**Authors:** George A. Yendewa, Gi-Ming Wang, Peter B. James, Samuel P.E. Massaquoi, Sahr A. Yendewa, Manal Ghazawi, Lawrence S. Babawo, Ponsiano Ocama, James B.W. Russell, Gibrilla F. Deen, Foday Sahr, Mustapha Kabba, Curtis Tatsuoka, Sulaiman Lakoh, Robert A. Salata

## Abstract

**Objective:** To estimate the prevalence and burden of chronic HBV in Sierra Leone.

**Methods:** We used electronic databases PubMed/MEDLINE, Embase, Scopus, ScienceDirect, Web of Science, Google Scholar and African Journals Online to systematically review articles reporting hepatitis B surface antigen (HBsAg) seroprevalence estimates in Serra Leone during 1997-2022. We estimated pooled HBV seroprevalence rates and assessed sources of heterogeneity

**Results:** Of 406 publications screened, 22 studies of total sample size 107,186 were included in the meta-analysis. The crude pooled HBV seroprevalence was 13.0% (95% CI 10.0-16.0) (I2=99%, p-heterogeneity<0.01), translating into 1.06 million people (95% uncertainty interval 0.81-1.30) or 1 in 8 Sierra Leoneans living with chronic HBV in 2021. Sensitivity analysis yielded a pooled HBV seroprevalence of 12.0% (95% CI 10.0-14.0) (sample size 104,968) (I2=98%, p-heterogeneity<0.001). The highest HBV seroprevalence estimates were among adolescents aged 10-17 years (17.0%, 95% CI 8.8-30.5), Ebola survivors (36.8%, 95% CI 26.2-48.8), people living with HIV (15.9%, 95% CI 10.6-23.0) and in the Northern (19.0%, 95% CI 6.4-44.7) and Southern (19.7%, 95% CI 10.9-32.8) regions. HBV seroprevalence progressively declined from 17.9% during 1997-2014 to 10.7% during 2020-2022.

**Conclusions:** These findings necessitate the urgent implementation of national HBV prevention and control programs in Sierra Leone.

## INTRODUCTION

Hepatitis B virus (HBV) is a double-stranded DNA virus of the *Hepadnaviridae* family that causes liver infection which can manifest as acute self-limiting hepatitis, fulminant liver failure, or chronic disease. Globally, an estimated 2 billion people—i.e., one-third of the world’s population— have been infected with HBV, with 1.5 million new cases occurring annually and 296 million chronically infected to date [1, 2]. The prevalence of HBV varies based on geographic location, with sub-Saharan African (SSA) and the West Pacific region accounting for 79% of chronic cases of HBV [1]. Despite the high global burden of the infection, current estimates suggest that only 10% of people living with HBV have been diagnosed [2]. Undiagnosed and untreated HBV infection confers an elevated life-time risk of liver-related complications including cirrhosis, hepatocellular carcinoma and end-stage liver disease events, which collectively accounted for 820,000 deaths in 2019 [1, 2].

Unlike the epidemic associated with the human immunodeficiency virus (HIV), viral hepatitis has comparatively received little public health policy focus until relatively recently. The Global Health Sector Strategy [3] and the Sustainable Development Goals [4] have recognized viral hepatitis as a neglected disease limiting the development of many low- and middle-income countries (LMICs) and have endorsed the elimination of HBV as a public health threat by the year 2030. As there is currently no curative therapy for HBV due to the persistence of the virus in hepatocytes, elimination efforts have focused on prevention through vaccination and treatment with antivirals. Vaccination is a potent public health strategy that is > 98% effective at preventing HBV infection [5], while treatment with nucleos(t)ide analogues (NA) can achieve durable suppression of viral replication and prevent liver-related morbidity and mortality [6, 7]. Despite the proven efficacy and cost-effectiveness of these strategies, many LMICs lack coordinated viral hepatitis prevention and control programs. In 2015, only three countries in the entire African region (viz., Algeria, Mauritania and Senegal) had well-outlined national action plans for combating HBV [8].

Sierra Leone is one of several countries in the West African region that is in the process of implementing a national prevention and control program for HBV infection. Accordingly, recent research efforts have focused on describing the characteristics of the HBV epidemic in the country. The majority of the studies—which have largely focused on blood donors, health workers, pregnant women, and people living with HIV (PWH)—have reported HBV seroprevalence rates ≥ 8% [10-15], consistent with the World Health Organization’s (WHO) classification for hyperendemicity [1]. Notwithstanding, there is limited understanding of the national prevalence, burden and overall impact of HBV due to insufficient surveillance systems. In hyperendemic settings such as Sierra Leone, HBV is most commonly acquired through vertical (mother-to-child) transmission or horizontal transmission during the early years of life [16]; yet antenatal screening remains limited in many such resource-constrained countries [17]. Of note, HBV vaccination was introduced in Sierra Leone in 2007 and incorporated into the Expanded Program on Immunization (EPI) in 2009 [18]; however, the birth-dose is yet to be implemented and full vaccine coverage for all age groups including vulnerable or high-risk groups such as pregnant women, healthcare workers, and people living with HIV has remained suboptimal [18].

To ensure successful HBV program implementation in Sierra Leone and aid ongoing efforts towards the 2030 HBV elimination goals, reliable estimations of the national prevalence and burden of HBV and identification of vulnerable or high-risk groups are essential for evidence-based policy planning and targeted public health interventions. In the absence of a national serosurvey, we performed a systemic review and meta-analysis of the available evidence to estimate the prevalence and burden of chronic HBV in Sierra Leone.

## METHODS

### Protocol registration

The study was prospectively registered with the International Prospective Register of Systematic Reviews (PROSPERO), registration number CRD42022337431. The conduct and reporting of the study was in accordance with the Preferred Reporting Items for Systematic Reviews and Meta-Analyses (PRISMA) guidelines (Supplementary File 1).

### Outcomes of the analysis

The primary outcomes of interest were the national prevalence and burden of chronic HBV infection in Sierra Leone, estimated as the pooled seroprevalence of the hepatitis B surface antigen (HBsAg) as detected by rapid diagnostic testing (RDT), enzyme-linked immunoassay (ELISA) and other approved tests in primary studies conducted in Sierra Leone. The secondary outcome of interest was the prevalence of chronic HBV in select populations in subgroup analysis. We defined two population groups, categorized as: (1) general populations, which included blood donors, healthcare workers, pregnant women seeking antenatal care, and other healthy individuals and (2) special populations with perceived higher risk of HBV infection, which consisted of PWH and Ebola virus disease (EVD) survivors.

### Country Characteristics

Sierra Leone is a country in West Africa with an estimated population of 8.14 million and a gross domestic product per capita of 516 United States in 2021 [19]. Since 2017, the country has been divided into 5 administrative regions, namely, the Northern, North West, Southern, and Eastern Provinces and the Western Area. For the purposes of this study, we retained the historical (pre-2017) classification by aggregating the contemporary Northern and North West Provinces as a single Northern region, the rationale being to simplify the process of determining study settings (Supplementary File 2). Freetown—the capital and largest city in Sierra Leone—is located in the Western Area and has an estimated population of 1.2 million. In recent years, Sierra Leone has faced a brutal civil war (1990-2001) and significant public health challenges, including parallel HIV and HBV epidemics and the West Africa Ebola epidemic (2014-2016), which have aligned to contribute to a fragile healthcare system [20]. In 2016, there were an estimated 1.4 doctors, nurses and midwives per 10,000 of the population, one of the lowest anywhere in the world [21]. The latest Sierra Leone Demographic Health Survey (2019) reported an infant mortality rate of 75 per 1,000 live births, maternal mortality of 717 deaths per 100,000 live births and adult mortality rate of 5.14 per 1,000 [22]. Despite these health indicators, the health expenditure per capita has remained low (i.e., 8.75% of the gross national product in 2020 [19], and over 60% of healthcare costs are financed by end-users through out-of-pocket payments [21].

### Study Design and Search Strategy

We systematically searched the online electronic databases PubMed/MEDLINE, Embase, Scopus, ScienceDirect, Web of Science, Google Scholar and African Journals Online for full-length texts of studies that assessed the seroprevalence of HBV in Serra Leone from inception to July 1, 2022. Our search terms included “Hepatitis B”, “Hepatitis B Virus”, “Hepatitis B infection” and “Sierra Leone” and were combined using the Boolean operators “OR” and “AND”. Additionally, we manually searched reference lists of studies to identify eligible articles.

### Inclusion and Exclusion Criteria

We considered full-text articles of observational studies published in English which reported the seroprevalence of HBsAg conducted in Sierra Leone from database inception to July 1st, 2022. Conference abstracts, case reports, case series, systematic reviews and meta-analysis, minireviews, editorials, commentaries, correspondence, studies that did not report the seroprevalence rate of HBsAg, studies with self-reported HBV infection status, and animal studies were not considered for inclusion. Where the full-text article of an otherwise eligible study was not available, the study was excluded. Additionally, we excluded studies conducted on Sierra Leonean-origin immigrant populations settled outside of the country.

### Study Selection, Screening and Data Extraction

Three authors (GAY, PBJ, and SPEM) independently screened the titles and abstracts of publications using the specified inclusion criteria and consulted others (RAS) in case of disagreements. After reaching consensus on eligibility of studies, all three authors (GAY, PBJ, and SPEM) independently extracted data from full-text articles and entered the information into a spreadsheet (Supplementary File 2). The following data were extracted: first author, year of publication, sampling year, study setting (region), study design, study population, sample size, gender, age, HBsAg prevalence, and diagnostic method used.

### Quality Assessment

Assessment of the methodological quality of the studies and risk of bias were undertaken by three authors (GAY, PBJ, and SPEM) using the Newcastle-Ottawa Scale (NOS) under the three domains of selection, comparability and exposure [22]. The NOS contains 9 items with a total maximum possible quality score (QS) of 9. Studies with QS of 9-8 were rated as very high quality, QS 7-6 as high quality, QS 5-4 as moderate quality, and QS 3-0 as low quality. However, we decided *a priori* to include all eligible studies regardless of quality rating.

### Data Analysis

The meta-analysis was performed using the “metaphor” and “dmetar” packages of the statistical software R (version 4.0.4) by pooling data within a random effects model using the DerSimonian and Laird estimator based on inverse variance weights. The magnitude of heterogeneity was reported as the heterogeneity (I2) index and its significance assessed by the χ2 test and Cochrane’s Q statistic. The degree of heterogeneity was reported as minimal (I2 < 25%), moderate (25% ≤ I2 < 50%) and high (I2 ≥ 50%). Subgroup and sensitivity analyses were performed to account for differences in HBV seroprevalence between studies. For subgroup analysis, subjects were categorized based on age, gender, geographic region, HIV status, EVD survivorship, healthcare workers, pregnancy, year of study and diagnostic method used. Publication bias was evaluated by both funnel plot asymmetry and Egger’s test. In all computations, differences were considered statistically significant at p-value < 0.05.

### Ethical approval

This study used data from published articles that are freely available in the public domain and therefore did not require ethical approval.

## RESULTS

### Identification and selection of studies

Figure 1 shows the PRISMA schematic flow of records identified. A total of 546 published records were found by literature search using PubMed/MEDLINE, Embase, Scopus, ScienceDirect, Google Scholar and African Journals Online. No additional studies were identified through searching of reference lists or other sources. After removing duplicates and screening studies based on title and abstract content, 28 articles were identified for full-text analysis. A total of 22 studies met the inclusion criteria and were included in the systematic review and meta-analysis.

**Figure 1.**
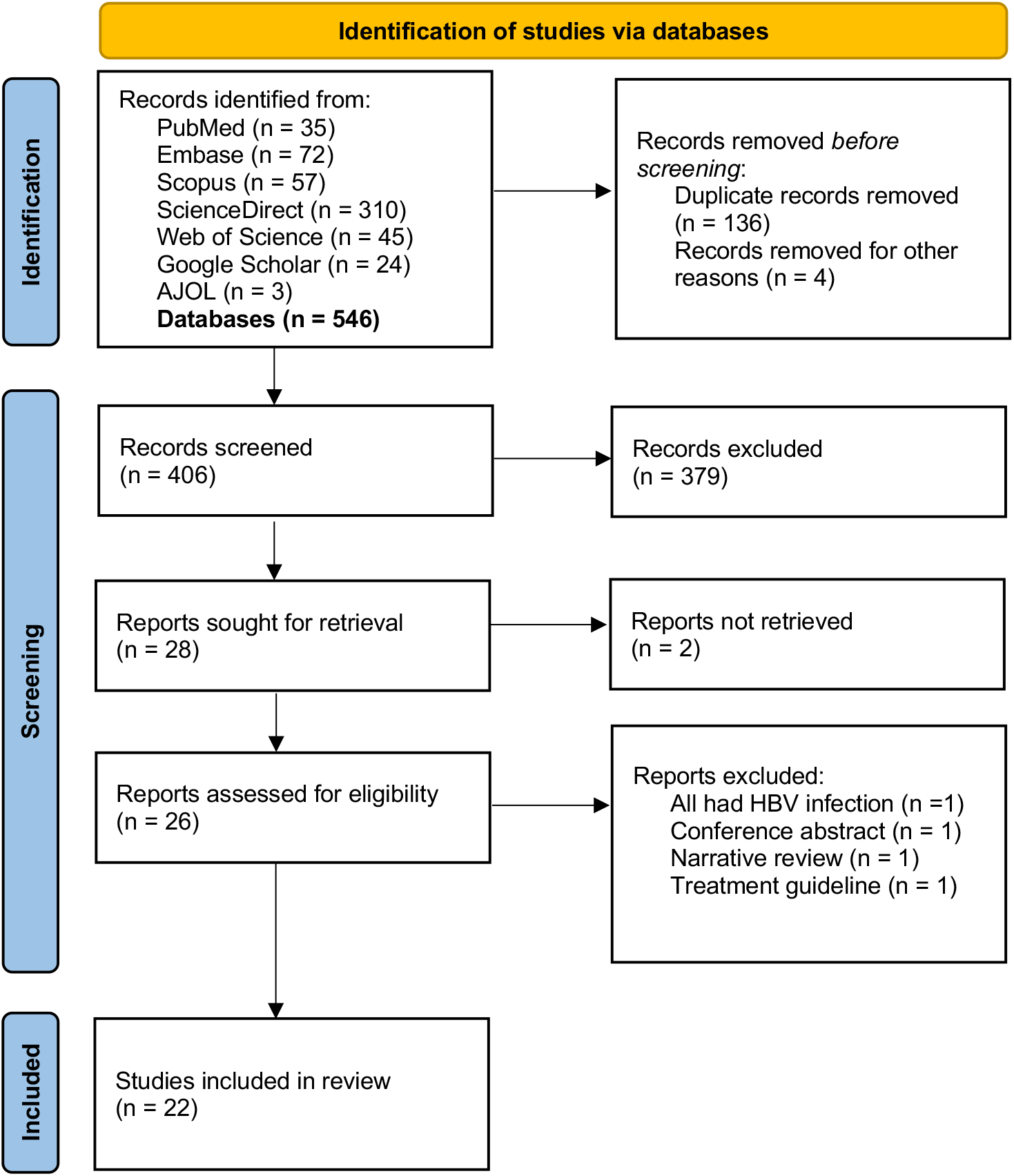
Flow diagram of systematic review, Sierra Leone 1997-2022

### Characteristics of included studies

Table 1 outlines the characteristics of the 22 studies included in the meta-analysis. The total sample size from the 22 studies included was 107,186. The individual study sample sizes ranged from 142 to 43,163. Based on the NOS for quality assessment, 6 (27%) studies were rated as very high quality (QS 9-8), 9 (41%) were of high quality (QS, 7-6), while the remaining 7 (32%) were of moderate quality (QS, 5-4). The studies included spanned a period of 25 years (1997-2022), with the majority (82%, 8) having been conducted after the incorporation of the HBV vaccine into the EPI schedule in Sierra Leone (i.e., 2010-2022).

**Table 1.**
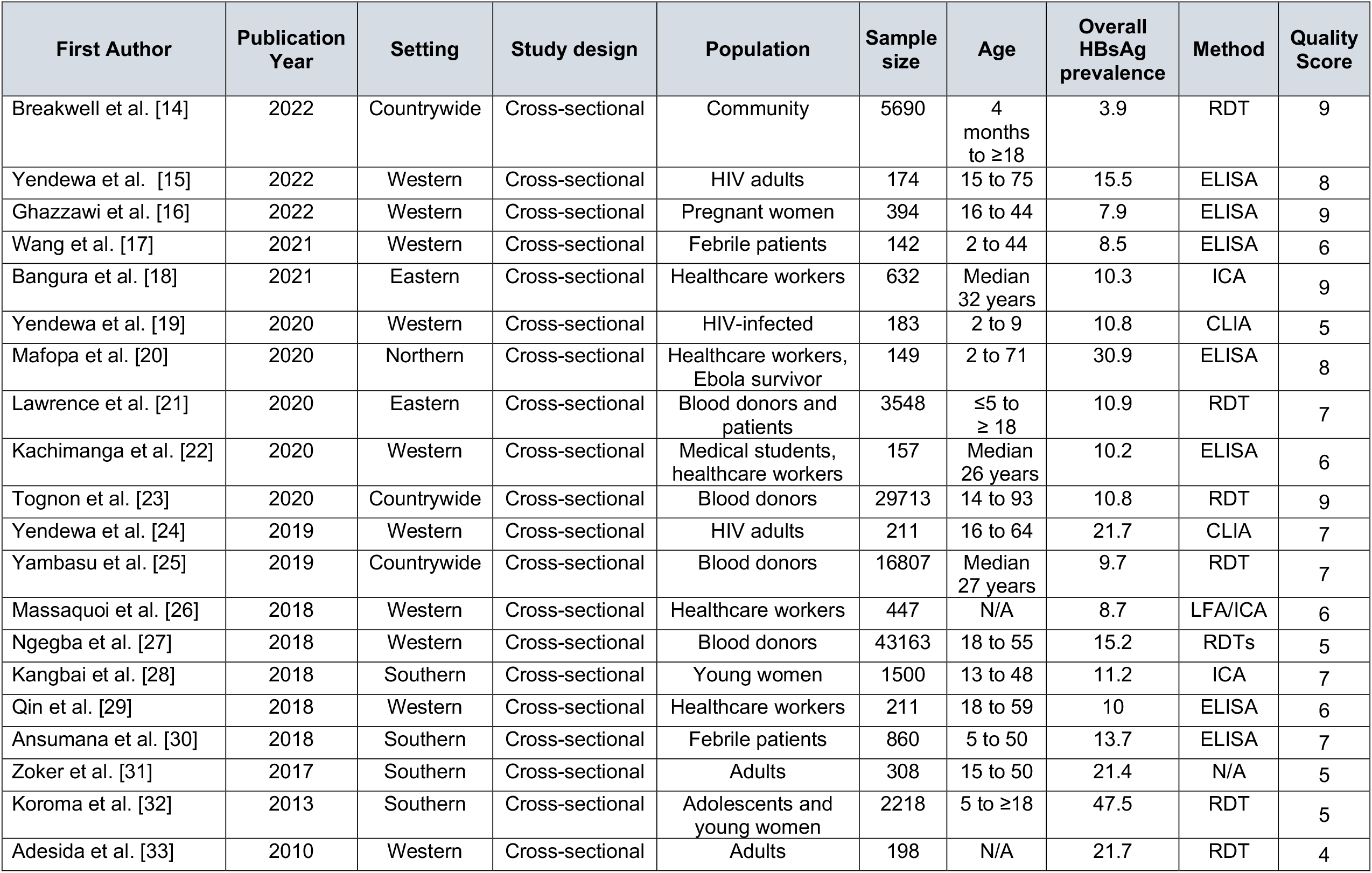
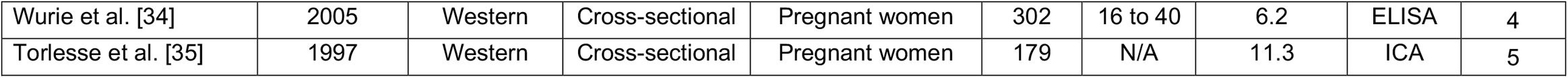
A summary of the descriptive characteristics of included studies

| First Author | Publication Year | Setting | Study design | Population | Sample size | Age | Overall HBsAg prevalence | Method | Quality Score |
| --- | --- | --- | --- | --- | --- | --- | --- | --- | --- |
| Breakwell et al. [14] | 2022 | Countrywide | Cross-sectional | Community | 5690 | 4 months to $\geq 18$ | 3.9 | RDT | 9 |
| Yendewa et al. [15] | 2022 | Western | Cross-sectional | HIV adults | 174 | 15 to 75 | 15.5 | ELISA | 8 |
| Ghazzawi et al. [16] | 2022 | Western | Cross-sectional | Pregnant women | 394 | 16 to 44 | 7.9 | ELISA | 9 |
| Wang et al. [17] | 2021 | Western | Cross-sectional | Febrile patients | 142 | 2 to 44 | 8.5 | ELISA | 6 |
| Bangura et al. [18] | 2021 | Eastern | Cross-sectional | Healthcare workers | 632 | Median 32 years | 10.3 | ICA | 9 |
| Yendewa et al. [19] | 2020 | Western | Cross-sectional | HIV-infected | 183 | 2 to 9 | 10.8 | CLIA | 5 |
| Mafofa et al. [20] | 2020 | Northern | Cross-sectional | Healthcare workers, Ebola survivor | 149 | 2 to 71 | 30.9 | ELISA | 8 |
| Lawrence et al. [21] | 2020 | Eastern | Cross-sectional | Blood donors and patients | 3548 | $\leq 5$ to $\geq 18$ | 10.9 | RDT | 7 |
| Kachimanga et al. [22] | 2020 | Western | Cross-sectional | Medical students, healthcare workers | 157 | Median 26 years | 10.2 | ELISA | 6 |
| Tognon et al. [23] | 2020 | Countrywide | Cross-sectional | Blood donors | 29713 | 14 to 93 | 10.8 | RDT | 9 |
| Yendewa et al. [24] | 2019 | Western | Cross-sectional | HIV adults | 211 | 16 to 64 | 21.7 | CLIA | 7 |
| Yambasu et al. [25] | 2019 | Countrywide | Cross-sectional | Blood donors | 16807 | Median 27 years | 9.7 | RDT | 7 |
| Massaquoi et al. [26] | 2018 | Western | Cross-sectional | Healthcare workers | 447 | N/A | 8.7 | LFA/ICA | 6 |
| Ngegba et al. [27] | 2018 | Western | Cross-sectional | Blood donors | 43163 | 18 to 55 | 15.2 | RDTs | 5 |
| Kangbai et al. [28] | 2018 | Southern | Cross-sectional | Young women | 1500 | 13 to 48 | 11.2 | ICA | 7 |
| Qin et al. [29] | 2018 | Western | Cross-sectional | Healthcare workers | 211 | 18 to 59 | 10 | ELISA | 6 |
| Ansumana et al. [30] | 2018 | Southern | Cross-sectional | Febrile patients | 860 | 5 to 50 | 13.7 | ELISA | 7 |
| Zoker et al. [31] | 2017 | Southern | Cross-sectional | Adults | 308 | 15 to 50 | 21.4 | N/A | 5 |
| Koroma et al. [32] | 2013 | Southern | Cross-sectional | Adolescents and young women | 2218 | 5 to $\geq 18$ | 47.5 | RDT | 5 |
| Adesida et al. [33] | 2010 | Western | Cross-sectional | Adults | 198 | N/A | 21.7 | RDT | 4 |
| Wurie et al. [34] | 2005 | Western | Cross-sectional | Pregnant women | 302 | 16 to 40 | 6.2 | ELISA | 4 |
| Torlesse et al. [35] | 1997 | Western | Cross-sectional | Pregnant women | 179 | N/A | 11.3 | ICA | 5 |

The prevalence of HBsAg ranged from 3.9% to 47.5% in the individual primary studies. The age of study participants ranged from 4 months to 75 years. All geographic regions of Sierra Leone were represented, with 12 (55%) studies from the Western Area, 4 (18%) from the Southern Province, 2 (9%) from the Eastern Province, 1 (5%) from the Northern and North West Provinces, 2 (9%) countrywide (Western, Eastern, Southern, Northern and North West) and 1 (5%) inter-regional (Western, Southern, Northern and North Western). The majority of the subjects were from the Western Area (55%), followed by Southern (18%), Eastern (13%) and Northern (13%) Provinces. Blood donors contributed the largest proportion of subjects (87%), followed by pregnant women (2.5%) and healthcare workers (1.4%); with children < 10 years, PWH, EVD survivors and others accounting for the remaining study participants (9%) (Table 1).

### Pooled prevalence and burden of HBV

Using a random-effects model, the crude pooled prevalence of chronic HBV in Sierra Leone was 13.0% (95% CI 10.0-16.0) in a pooled sample of 107,186 individuals (I2 = 99%, p-heterogeneity < 0.01) (Figure 2). Using the 2021 national population estimate of 8.14 million [19] yielded an estimated national chronic HBV burden of 1.06 million infected people (95% uncertainty interval 0.81-1.30). In sensitivity analysis, we removed the study with the highest seroprevalence of HBV (Koroma et al., 2013) to ensure stability of the crude prevalence estimates. This yielded a sensitive HBV seroprevalence of 12.0% (95% CI 10.0%-14.0%) in a pooled sample of 104,968 individuals (I2= 98%, p-heterogeneity < 0.001), which did not significantly differ from the crude estimates (Supplementary File 3).

**Figure 2.**
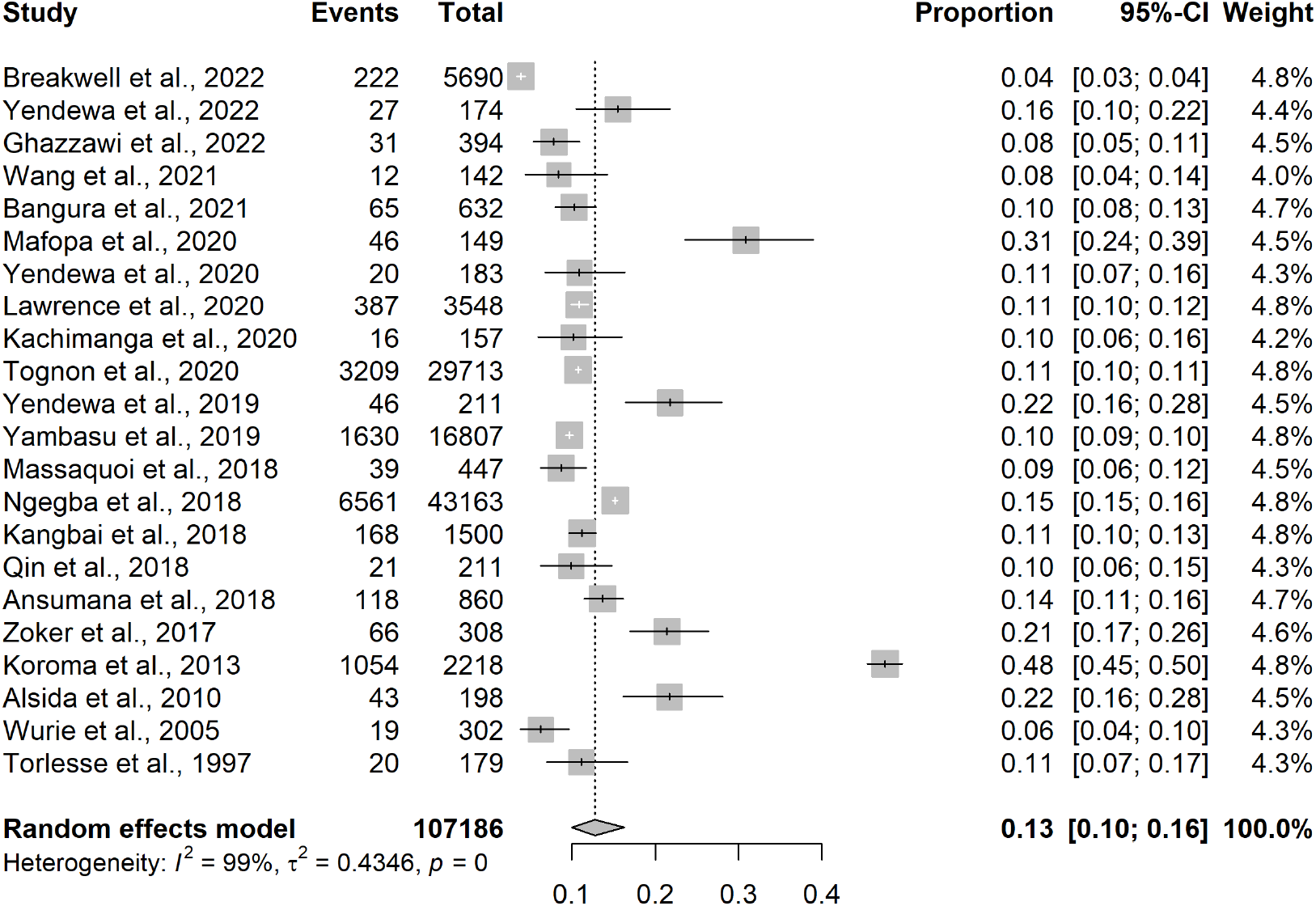
Forest plot of the pooled seroprevalence of HBV in Sierra Leone, 1997-2022

### Subgroup analysis

The results of the subgroup analysis are shown in Table 2. Based on the studies which provided age-specific estimates, adolescents aged 10-17 years had the highest HBV seroprevalence (17.0%, 95% CI 8.8-30.5), followed by adults aged ≥ 18 years (12.4%, 95% CI 9.9-15.4). Children aged 5-9 years had the lowest HBV seroprevalence rates (1.6%, 95% CI 1.1-2.2), while under-fives had HBV seroprevalence of 2.9% (95% CI 0.6-12.6). Females had a slightly higher HBV seroprevalence (13.4%, 95% CI 7.4-23.1) than their male counterparts (10.5%, 95% CI 7.2-15.0). The Northern (19.0%, 95% CI 6.4-44.7) and Southern (19.7%, 95% CI 10.9-32.8) regions had higher HBV seroprevalence rates compared with the Western Area (11.2%, 95% CI 8.9-14.1) and Eastern Province (10.1%, 95% CI 9.1-11.2) (Figure 3). Regarding general populations, the HBV seroprevalence among healthcare workers was 11.9% (95% CI 7.7-18.0), while pregnant women seeking antenatal care had a HBV seroprevalence of 9.7% (7.7-14.6). Within the two special population groups, EVD survivors had higher HBV seroprevalence (36.8%, 95% CI 26.2-48.8), while HIV-infected individuals had an HBV seroprevalence estimate of 15.9% (95% CI 10.6-23.1). Of the testing methods used, RDT accounted for the highest seroprevalence rates (13.9%, 95% CI 7.4-24.8) versus ELISA (11.8% 95% CI 8.1, 16.8) and other diagnostic methods (13%, 95% CI 9.9-17.2). Over the 25-year period covered by the study, there was a trend towards lower HBV seroprevalence rates from 17.9% (95% CI 6.7-39.8) estimated from the studies conducted before 2015, 13.3% (95% CI 10.4-16.9) during 2015-2019 and 10.7% (95% CI 7.5-14.9) estimated from the studies published during 2020-2022. Overall, heterogeneity remained high among studies, with the sources of heterogeneity significantly accounted for by age and EVD survivorship (p-difference < 0.001, respectively). Geographic regions contributed substantially to HBV seroprevalence heterogeneity but did not attain statistical significance (p-difference 0.097) (Table 3).

**Table 2.** Subgroup analysis of the HBV seroprevalence estimation in Sierra Leone, 1997-2022

| Subgroup | Variable | Number of studies | Prevalence % (95% CI) | $I^2$ % | p-Value for difference |
| --- | --- | --- | --- | --- | --- |
| **Age** | < 5 | 2 | 2.9 (0.6, 12.6) | 96.0 | **< 0.001** |
|  | 5 to 9 | 1 | 1.6 (1.1, 2.2) | NA |  |
|  | 10 to 17 | 1 | 17.0 (8.8, 30.5) | NA |  |
|  | ≥ 18 | 11 | 12.4 (9.9, 15.4) | 97.3 |  |
| **Sex** | Males | 17 | 10.5 (7.2, 15.0) | 98.0 | 0.477 |
|  | Females | 7 | 13.4 (7.4, 23.1) | 99.4 |  |
| **Geographic region** | Western | 13 | 11.2 (8.9, 14.1) | 96.2 | 0.097 |
|  | Eastern | 3 | 10.1 (9.1, 11.2) | 66.5 |  |
|  | Northern | 2 | 19.0 (6.4, 44.7) | 97.9 |  |
|  | Southern | 5 | 19.7 (10.9, 32.8) | 99.6 |  |
| **Study population** | HIV positive | 3 | 15.9 (10.6, 23.1) | 76.1 | 0.320 |
|  | Ebola survivor | 1 | 36.8 (26.2, 48.8) | NA | **< 0.001** |
|  | Healthcare worker | 4 | 11.9 (7.7, 18.0) | 79.8 | 0.632 |
|  | Pregnant women | 4 | 9.7 (6.3, 14.6) | 68.6 | 0.210 |
| **Diagnostic test** | ELISA | 8 | 11.8 (8.1, 16.8) | 89.3 | 0.867 |
|  | RDT | 7 | 13.9 (7.4, 24.8) | 99.8 |  |
|  | Others | 7 | 13.0 (9.8, 17.2) | 87.8 |  |
| **Year of study** | < 2015 | 4 | 17.9 (6.7, 39.8) | 98.6 | 0.434 |
|  | 2015-2019 | 8 | 13.3 (10.4, 16.9) | 98.0 |  |
|  | ≥ 2020 | 10 | 10.7 (7.5, 14.9) | 97.1 |  |

**Figure 3.**
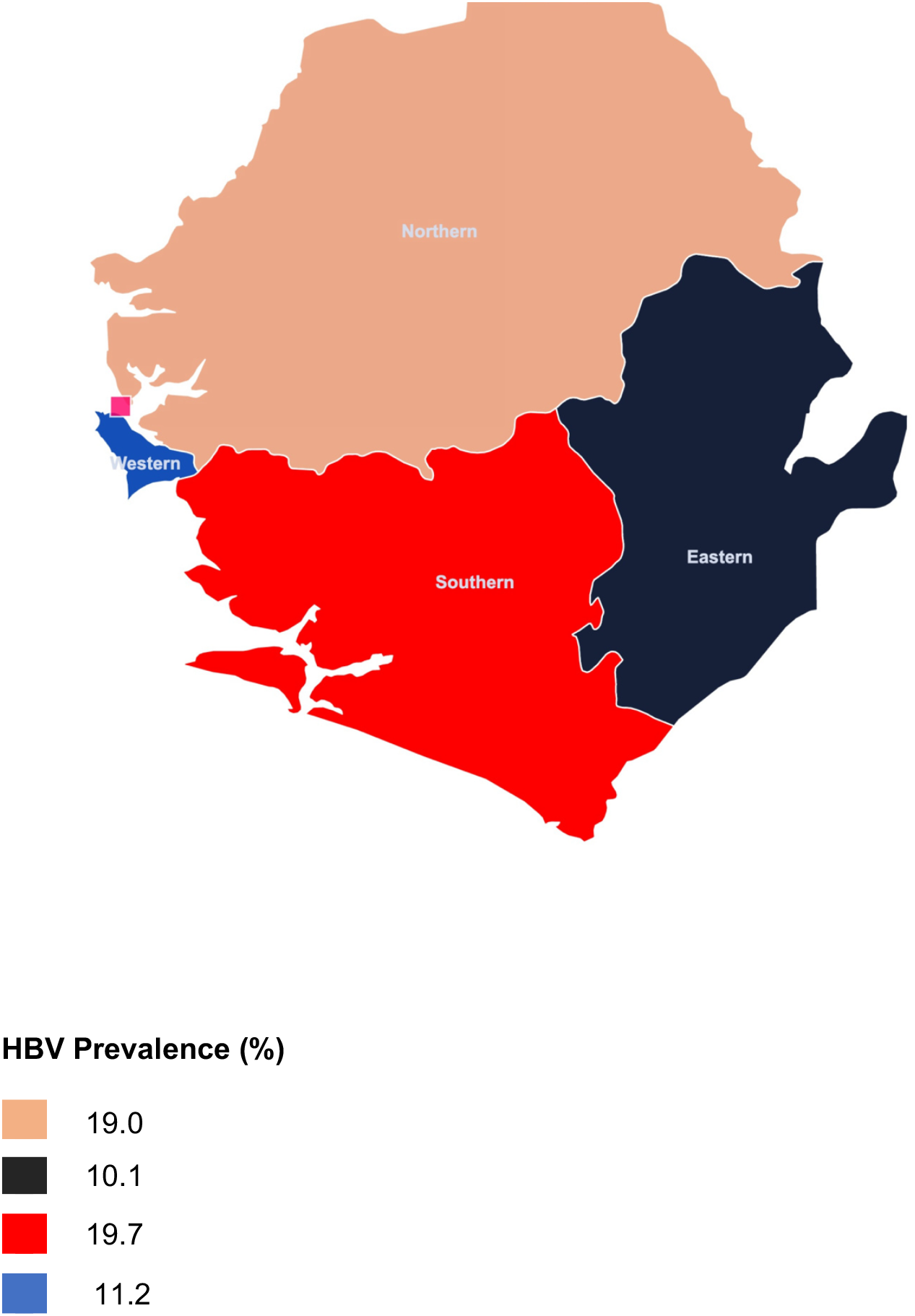
Map of Sierra Leone with estimated HBV prevalence by region (not drawn to scale)

**Figure 3.**
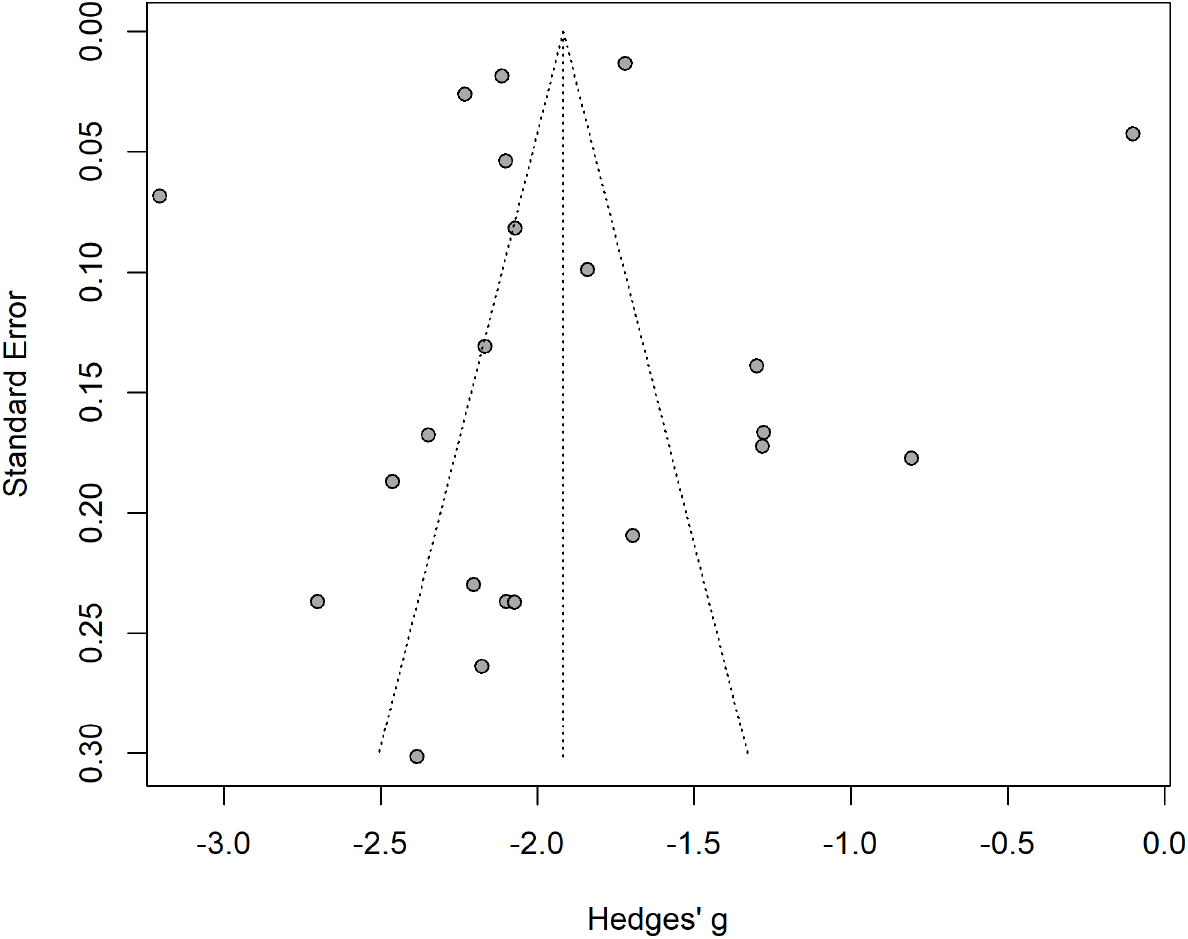
Funnel plot of the seroprevalence of HBV estimation in Sierra Leone, 1997-2022

### Publication bias

Figure 3 shows the funnel plot. The Egger’s regression did not show asymmetry (p = 0.892), indicating no evidence of publication bias.

## DISCUSSION

This is the first systematic review and meta-analysis to estimate the national prevalence and burden of chronic HBV in Sierra Leone. The overall goal was to synthesize evidence, to help inform ongoing efforts seeking to combat the HBV epidemic in the country. Data was pooled from general and special (high-risk) populations and all geographic regions were represented. The crude pooled prevalence of chronic HBV was 13%, which did not significantly differ from the 12% estimate obtained in sensitivity analysis. Based on the 2021 country population estimates, about 1.06 million people or 1 in every 8 Sierra Leoneans are living with chronic HBV infection. Most (68%) of the primary studies included in the analysis were rated as having high methodological quality, suggesting a reliable estimate. Our findings are consistent with recent data from Ghana [23], Nigeria [24], Cameroon [25] and other countries in West Africa [26], which have recorded HBV prevalence rates ≥ 8%, in line with WHO’s threshold for hyperendemicity [1].

We observed a trend towards decreasing HBV prevalence in Sierra Leone over the 25-year period covered by the study. The pooled HBV prevalence was 17.9% during 1997-2014, 13.3% during 2015-2019 and 10.7% from 2020-2022. This is encouraging and suggests that the national immunization program implemented in 2009 is having a positive impact on the HBV epidemic. According to UNICEF estimates, the HBV vaccine coverage for infants in Sierra Leone was 92% in 2020, surpassing the estimated global coverage of 80% [27]. This is reflected in the low HBV seroprevalence observed among vaccinated under-fives (2.9%) and children aged 5-9 years (1.6%) in subgroup analysis. As in many LMICs, the pentavalent vaccine is the form in which HBV immunization is currently available in Sierra Leone. Under the national immunization program, the pentavalent vaccine is administered at 6, 10 and 14 weeks after birth. However, as previously noted, the birth-dose, which is administered within the first 24 hours postpartum, is yet to be incorporated into the immunization schedule. Additionally, HBV prevalence among pregnant women has remained high (9.7%) (REF)? while vaccination coverage has remained extremely low among this group and other adults born before vaccination was made available. Consequently, the risk of mother-to-child transmission remains high, emphasizing the need for implementation of the birth-dose, scaling up of antenatal screening, and vaccination for susceptible individuals.

We did not find significant differences in HBV seroprevalence based on sex, pregnancy status, being a healthcare worker, or testing method, as some studies from SSA have previously described [Ref]. However, HBV prevalence among adolescents aged 10-17 years was 17%, which was higher than in any other age category. Data on the prevalence and impact of HBV infection on adolescents is scarce; however, recent studies from West Africa and elsewhere suggest that adolescents and young adults are a high-risk group that has received little attention in the global elimination efforts [Ref]. A meta-analysis by Abesig *et al* [Ref] estimated a high HBV prevalence (14.3%) among adolescents in Ghana (2015-2019) compared to a prevalence of 8.36% among adults. In a 2018 study from a clinic screening for sexually transmitted infections (STIs) in Nigeria, Nejo *et al* [Ref] found a low HBV seroprevalence among adolescents (1.9%); however, the risk of HBV was 9-fold higher (HBV seroprevalence 17%) in sexually actively adolescents.

A plausible explanation for the disproportionately high impact of HBV on adolescents and young adults in SSA is that this group tends to have a high burden of risk factors that increase susceptibility to horizontal transmission of HBV and other STIs (notably HIV), including intravenous drug use, multiple sexual partners, tattooing and other body scarification procedures, and insufficient knowledge of disease processes due to cultural norms that restrict discussions around sexual practices [Ref]. Others have noted that in endemic settings in SSA, most adolescents with chronic HBV are overlooked as they predominantly present in the immunotolerant phase, which is characterized high rates of viral replication without evidence of hepatic inflammation and therefore rarely require treatment [Ref]. This highlights the need for further research into prevention, screening and treatment strategies targeted at this population.

Our study uncovered regional disparities in HBV seroprevalence in Sierra Leone. The Northern and Southern regions had similarly high HBV seroprevalence rates (19% and 19.7% respectively), almost double the rates in the Western Area (11.2%) and Eastern region (10.1%). The reasons for these regional variations are unclear but may include disparities in antenatal screening, uptake of vaccination and availability of treatment services [Ref]. Other barriers hampering HBV control efforts that have been documented in SSA include low socio-economic status, poor knowledge of HBV and low health literacy in general [Ref]. Emerging research also suggests that similar to HIV, HBV-related stigma is pervasive in communities across SSA and may be influencing health-seeking behaviors, but this phenomenon is poorly understood [Ref]. These factors need to be further explored, as understanding the reasons behind these regional differences may be crucial to designing evidence-based healthcare strengthening strategies to effectively tackle the HBV epidemic in the country.

We further observed a higher HBV seroprevalence among HIV-infected individuals (15.9%). HIV is a well-recognized risk factor for HBV infection and *vice versa*, due to shared risk factors and routes of transmission [Ref]. A prominent feature of the HIV epidemic in West Africa is its intersection with HBV, which is a major driver of disease progression and outcomes in this region [16]. A recent meta-analysis by McGowan et al [Ref] estimated a global HBV seroprevalence of 7.6% among PWH. The highest HIV/HBV coinfection prevalence was observed in West and Central Africa at 16.4%, more than 2-fold higher than the global prevalence rate [25], which is consistent with our findings. The consequences of HIV/HBV co-infection have been well-documented and include faster progression to the acquired immunodeficiency syndrome (AIDS), liver cirrhosis and hepatocellular carcinoma [Ref]. Thus, in HBV-hyperendemic countries such as Sierra Leone, successfully tackling the HIV epidemic will also require incorporating parallel efforts aimed at simultaneously combating HBV, in order to meet the 2030 global HIV and HBV elimination targets. This warrants greater integration of HIV and HBV services, especially in West Africa which is disproportionately affected by the HBV epidemic.

The prevalence of chronic HBV among EVD survivors was 36.8%. This estimate was from a single study by Mofopa et al [Ref], which makes it difficult to draw generalizable inferences to the wider population. The West Africa Ebola epidemic of 2014-2016 was a major public health crisis that resulted in 30,000 symptomatic cases and over 11,000 EBV-related deaths in Sierra Leone, Guinea and Liberia [Ref]. Despite the large number of EVD survivors in the region, little is known about how EBOV may be interacting with endemic viruses such as HBV and HIV co-circulating in the general population in West Africa and other EBOV-endemic regions in SSA. Of note, the study by Mofopa et al [Ref] was conducted in the Northern Province, which had high HBV seroprevalence at baseline (19%), which could partner explaining this finding. More research is needed to investigate the potential links between EBOV, HBV and other endemic viruses and potential impact on disease pathogenesis and outcomes in this setting.

## CONCLUSIONS

In conclusion, we observed a high prevalence (13%) and burden of chronic HBV in Sierra Leone, four decades after the first HBV vaccine was produced, despite a progressive downward trend in seroprevalence in recent years. Adolescents, Ebola survivorship, PWH and residing in the Northern and Southern regions were major predisposing factors to having chronic HBV in this setting. These findings necessitate the urgent implementation of national HBV prevention and control programs in Sierra Leone to meet the 2030 viral hepatitis elimination goals.

## DATA STATEMENT

All the data extracted from studies are contained in the manuscript and supplemental materials.

## AUTHOR CONTRIBUTIONS

G.A and R.A.S. conceptualized the study, with input from GW, CT, SAY, and SL. Literature search, study screening and selection of studies was performed by GAY, PBJ and SPEM. GW and CT conducted the statistical analyses. GAY drafted the first manuscript version, with contributions from GW, PBJ, SAY, LSB, PO, SL and RAS. All authors interpreted the data, revised the manuscript and contributed important intellectual content. All agree with the results and conclusions of this article.

